# Rapid implementation of cross-sectional study: Post-acute sequelae of SARS-CoV-2 (PASC) in a racially and ethnically diverse sample in Illinois

**DOI:** 10.1101/2021.04.29.21256304

**Authors:** Robin J. Mermelstein, Meenakshy Aiyer, Christine Canfield, David Chestek, Judith A. Cook, Marina Del Rios, Kathleen R. Diviak, Angela M. Ellison, Howard S. Gordon, Sai Dheeraj Illendula, Manasa Kandula, Jonathan D. Klein, Karen Larimer, James Lash, Janet Lin, Jeffrey A. Loeb, Teresa J. Lynch, Hugh Musick, Richard M. Novak, Heather M. Prendergast, Jerry A. Krishnan

## Abstract

Little is known about the pattern and course of recovery following acute COVID-19. Increasing numbers of reports describe persistent illness following infection with SARS-COV-2, also known as Post-Acute Sequelae of SARS-CoV-2 (PASC). This report describes the methods and results of a multi-pronged strategy to rapidly identify and enroll, over a one week period in April 2021, a racially and ethnically diverse sample of individuals and to characterize PASC among a this diverse sample. Participants were recruited through community outreach, clinical registries, and research registries across four cities in Illinois to complete an online survey. We examined presence of symptoms among 246 individuals who were at least three months past testing positive for SARS-CoV-2. Respondents were 70% female; 48% Hispanic/Latinx; 18% Black, and 28% White. Most had mild illness (78% were not hospitalized), and 26% reported they had not yet returned to their usual health within 3 months of their diagnosis. The most prevalent symptoms persisting 3-months following COVID-19 diagnosis included fatigue (20%), difficulty thinking (19%), problems with taste or smell (15%), and muscle or body aches (15%). In a multivariable logistic regression model, older age (40-59 vs. 18-39 years: adjusted odds ratio [aOR] = 0.46 [95% confidence interval, 0.24 to 0.90]) and having been hospitalized with COVID-19 (vs. not hospitalized: aOR = 0.28 [0.12 to 0.64]) were independently associated with a lower likelihood of recovery within 3 months. Compromised health continued well beyond the acute phase of COVID-19 in our ethnically diverse sample, especially among older individuals and those who were hospitalized. The partnerships with community- and faith-based organizations developed for the current study offer the potential to broadly disseminate study findings and to further understand and mitigate underlying determinants of risk, severity, and duration of PASC.

## INTRODUCTION

Coronavirus disease 2019 (COVID-19) is caused by a novel coronavirus, SARS-CoV-2. Efforts to end the COVID-19 pandemic have focused largely on the early phases of the natural history of the disease: defining and implementing evidence-based practices for the prevention of SARS-CoV-2 infection and treatment of acute sequelae.(1-3) Much less is known about the patterns and time course of recovery following the acute stages of infection and illness. While efforts directed toward prevention and clinical management continue, an increasing number of reports have described the potential for persistent illness following infection with SARS-CoV-2, also known as Long COVID, Post-Acute Sequelae of COVID, or Post-Acute Sequelae of SARS-CoV-2 (PASC). To address gaps in our understanding about the causes and ultimately the prevention and treatment of the full course PASC, the National Institutes of Health (NIH) recently announced the launch of the trans-NIH PASC Initiative.(4)

Most published reports about PASC focus on hospitalized patients with COVID-19, even though more than 90% of SARS-CoV-2 illnesses have no symptoms or result in mild symptoms and are managed as outpatients.(5-9). In the U.S., Hispanic/Latinx and Black populations have a higher risk of SARS-CoV-2 infection, as well as COVID-19 associated hospitalizations and death.(10) More recent studies on PASC in the U.S. have included participants across the range of COVID-19 disease severity(11-14), but include low proportions of racial or ethnic minorities.

A better understanding of the risks and characteristics of PASC by race, ethnicity and severity of COVID-19 can help to tailor public health messaging about the consequences of COVID-19 and the importance of participating in research that will define best practices for the prevention and management of PASC. As the COVID-19 pandemic evolves, approaches to quickly sample high-risk populations are needed for efficient and effective public health initiatives. The Illinois Network for Post-Acute Sequelae of SARS-CoV-2 (ILLInet-PASC) is a community-engaged collaboration between community- and faith-based organizations, public health departments, and health systems in metropolitan and surrounding areas of Chicago, Peoria, Rockford, and Urbana, Illinois.(15) The objectives of this report are to describe the methods and results of a multi-pronged strategy to rapidly (over a 1-week period) identify and enroll a diverse sample, and to characterize PASC in a predominantly Hispanic/Latinx and Black population in Illinois.

## METHODS

### Overview of Design and Recruitment

This study followed the Strengthening the Reporting of Observational Studies in Epidemiology (STROBE) guidelines. We employed a cross-sectional study design to collect self-reported information using an on-line questionnaire programmed with REDCap from April 7 to 12, 2021.(16) Our goal was to simultaneously employ a set of rapid recruitment approaches to maximize enrollment within a 7-day window for data collection that would yield a diverse sample of individuals living in Illinois who had tested positive for SARS-CoV-2 and who were willing to share their recovery experiences. To ensure rapid launch of the study protocol, we leveraged existing institutional review board (IRB)-approved research studies for which investigators had ongoing approval to contact participants for potential study recruitment; partnerships with the Chicago Department of Public Health and Peoria City & County Health Department (PCCHD); and on-going collaborations with community-based and faith-based organizations through the University of Illinois at Chicago (UIC) Office of Community Engagement and Neighborhood Health Partnerships (OCEAN-HP), Mile Square Federally Qualified Health Centers (Mile Square FQHC), UIC Department of Pediatrics (CHECK program), UIC School of Public Health, the UIC College of Medicine in Chicago, Peoria, and Rockford. IRBs in Chicago (Protocol # 2021-0410), Peoria (Protocol #1745454-1), Urbana (Protocol # #21CRU3374), and Jesse Brwon VA Medical Center (Protocol #1621031-1) reviewed the ILLInet-PASC study protocol and determined the study to be exempt from further review.

There were three sources of study participants.

1. *Community outreach*. We provided the IRB-approved ILLInet-PASC flyers or the URL or QR code for the on-line survey to community-based and faith-based partner organizations. Community partners used a variety of approaches, including email and telephone-based interviews of individuals to complete on-line survey. The on-line surveys were available in both English and Spanish.
2. *COVID-19 research registries*. The investigator of on-going COVID-19 studies sent an email invitation to potential participants describing the study. Individuals could access the link in the email to learn more and complete the survey if interested.
3. *COVID-19 clinical registries*. The investigator with controlling access to these registries sent an email with information about the study and survey to the registry participant. Individuals from the registry could access the link in the email to learn more and complete the survey if interested.

The on-line REDCap survey, in English and Spanish, was designed to take no more than 10 minutes to complete. Only adults who self-reported an age of 18 years or older were eligible to complete the survey. Once participants submitted the survey, they were invited to receive a $10 Amazon^©^ gift card (e-code). To receive the Amazon^©^ e-code, participants were directed to a second REDCap survey to provide their contact information, separating data from identifying information.

### Questionnaire domains

We assessed four domains.

1. *Demographics*. Participants were asked about their age, gender, race and ethnicity.
2. *Comorbidities*. Participants were asked to report health conditions diagnosed by their healthcare provider using a list of 18 potential comorbidities. Participants were also asked whether they currently smoked tobacco or vaped nicotine or cannabis.
3. *COVID-19 testing, results, and severity*. Participants were asked to indicate the month(s) they were tested for COVID-19 (from January, 2020 – April, 2021), and any month(s) they had a positive test result for COVID-19. Multiple months could be selected. Severity of acute illness from COVID-19 was defined on the basis of the level of healthcare received: mild (none, outpatient, or emergency department [ED]), moderate (hospitalization without intensive care unit), and severe (hospitalization with intensive care unit [ICU]).(17)
4. *Current symptoms and timeline for recovery*. Participants were asked if they currently experienced symptoms from a list of 26 possible symptoms based on previously published studies. (18,21) For each current symptom, participants indicated whether it was better, worse, or unchanged compared to before they were diagnosed with COVID-19. Participants were also asked about whether they had recovered to their usual state of health and asked to select: No; Yes, recovered within 2 weeks; Yes, recovered within 1 month; Yes, recovered within more than 1 month, but less than 2 months; Yes, recovered within more than 2 months, but less than 3 months; Yes, recovered after 3 months. We considered responses of “No” and “recovered after 3 months” as not returning to baseline health by 3 months (PASC-3 months).

### Analyses

To ensure adequate follow-up time to evaluate the risk of PASC-3 months, analyses focused on individuals who reported testing positive for COVID-19 at least 3 months prior to survey completion. We used descriptive statistics of frequencies of responses, as well as chi-square analyses to examine differences in frequencies of symptoms by subgroups (e.g., by gender, race/ethnicity). Multivariable logistic regression analysis was used to determine if demographics (age, gender, race/ethnicity) and severity of COVID-19 (mild, moderate or severe) were independent predictors of incomplete return to usual health by 3 months post-COVID-19 diagnosis. A significant difference was defined as a 2-tailed p-value<0.05.

## RESULTS

Of 401 participants who completed the on-line survey from April 7 to 12, 2021, the majority were recruited from community outreach sources (N=260, 65%), followed by COVID-19 research registries (N=75, 19%) and COVID-19 clinical registries (N=66, 16%). Most of the 401 participants were Hispanic/Latinx (47%) or Black/African-American (20%). A total of 284 (71%) participants reported a positive COVID-19 test, with a median of 4 months prior to completing the survey (range 1 to 15 months). Of these 284, 246 (86.6%) completed the survey 3 or more months after a positive COVID-19 test (Table 1).

**Table 1:**
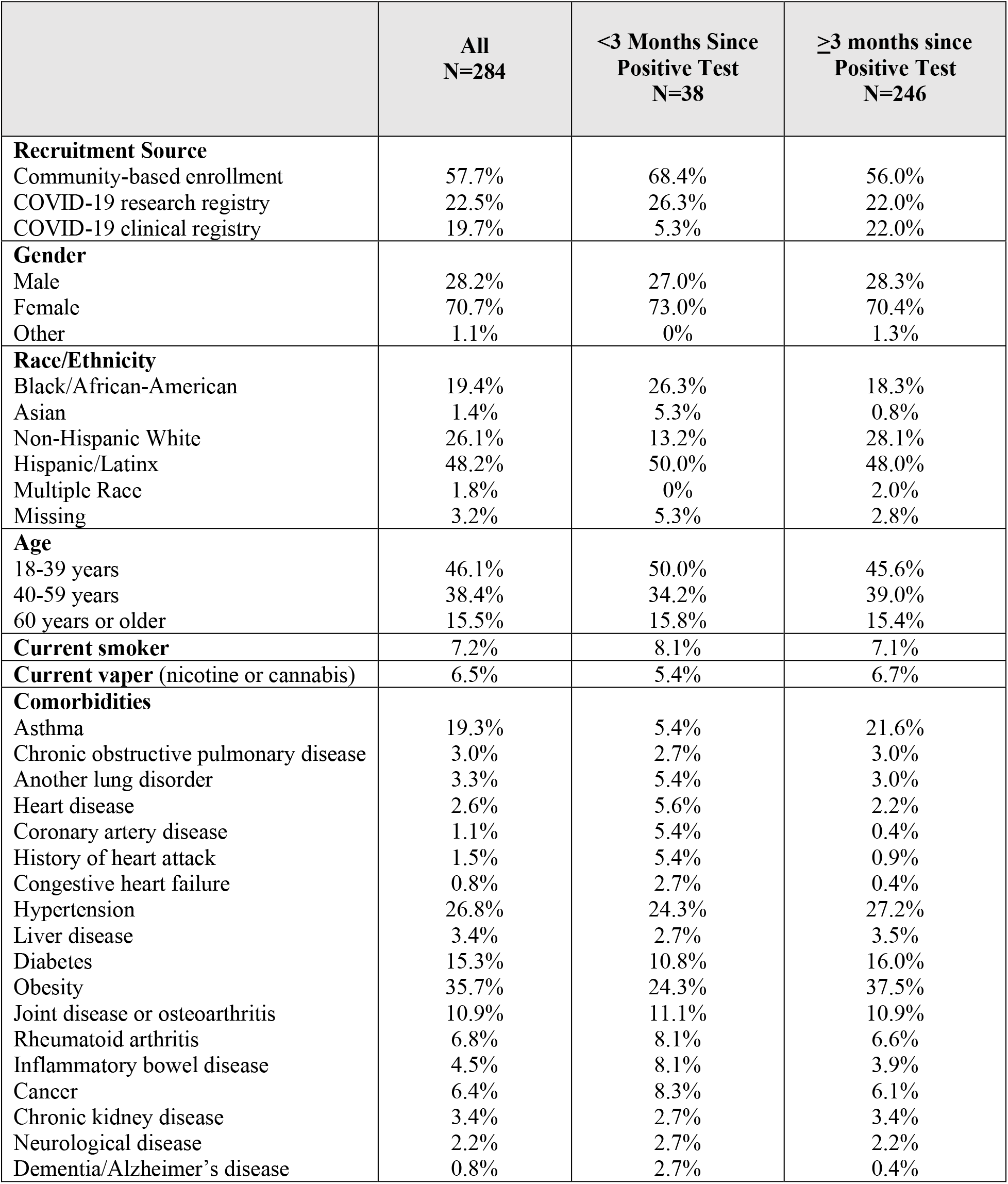
Characteristics of participants with a positive COVID-19 test.

The majority of these 246 participants (85%) had mild COVID-19, defined as not seeking medical care (53%), receiving only outpatient care (25%), or receiving care in the Emergency Department (7%). An additional 9% had moderate COVID-19, defined as being hospitalized without ICU-level care; and 5% had severe COVID-19, defined as hospitalized with ICU-level care. Most were from community-based sources (56%), women (70%) and at least 40 years old (54%). Over two-thirds were non-White (48% Hispanic/Latinx, 18% Black, 2% multiple races, 1% Asian). The five most prevalent comorbidities were obesity (38%), hypertension (27%), asthma (22%), diabetes (16%), and joint disease or osteoarthritis (11%). About 7% reported current smoking; a similar proportion (7%) reported current vaping.

### Current symptoms and recovery timeline

Three or more months after the diagnosis of COVID-19, about 1 in 5 reported fatigue (20%), trouble concentrating (18%), or trouble thinking (18%) that was worse than before they were diagnosed with COVID-19 (Table 2). About 1 in 4 (26%, 75/284) indicated they had not recovered fully (Table 3). Incomplete recovery at 3 months was significantly more likely among those who were older and among those with moderate or severe COVID-19.

**Table 2:** Current symptoms that are worse since COVID-19 diagnosis, among participants >3 months since positive test (N=246)

| Symptom | Prevalence |
| --- | --- |
| Fatigue | 20.3% |
| Difficulty thinking | 18.8% |
| Trouble concentrating | 18.3% |
| Body or muscle aches | 15.2% |
| Problems with taste or smell | 15.4% |
| Trouble sleeping | 15.4% |
| Feeling anxious | 13.9% |
| Trouble breathing/shortness of breath | 14.4% |
| Feeling depressed | 12.2% |
| Weakness in arms or legs | 10.9% |
| Headache | 10.4% |
| Decrease in appetite | 7.8% |
| Dizziness | 7.4% |
| Painful toes or fingers | 7.4% |
| Cough | 5.6% |
| Chest pain | 6.5% |
| Stuffy nose or congestion | 5.2% |
| Nausea or vomiting | 3.9% |
| Change in color of toes or fingers | 3.5% |
| Sore throat | 2.6% |
| Swollen leg | 2.6% |
| Pain in stomach or abdomen | 2.6% |
| Diarrhea | 2.2% |
| Rash | 1.7% |
| Chills | 1.3% |
| Fever | 1.3% |

**Table 3:**
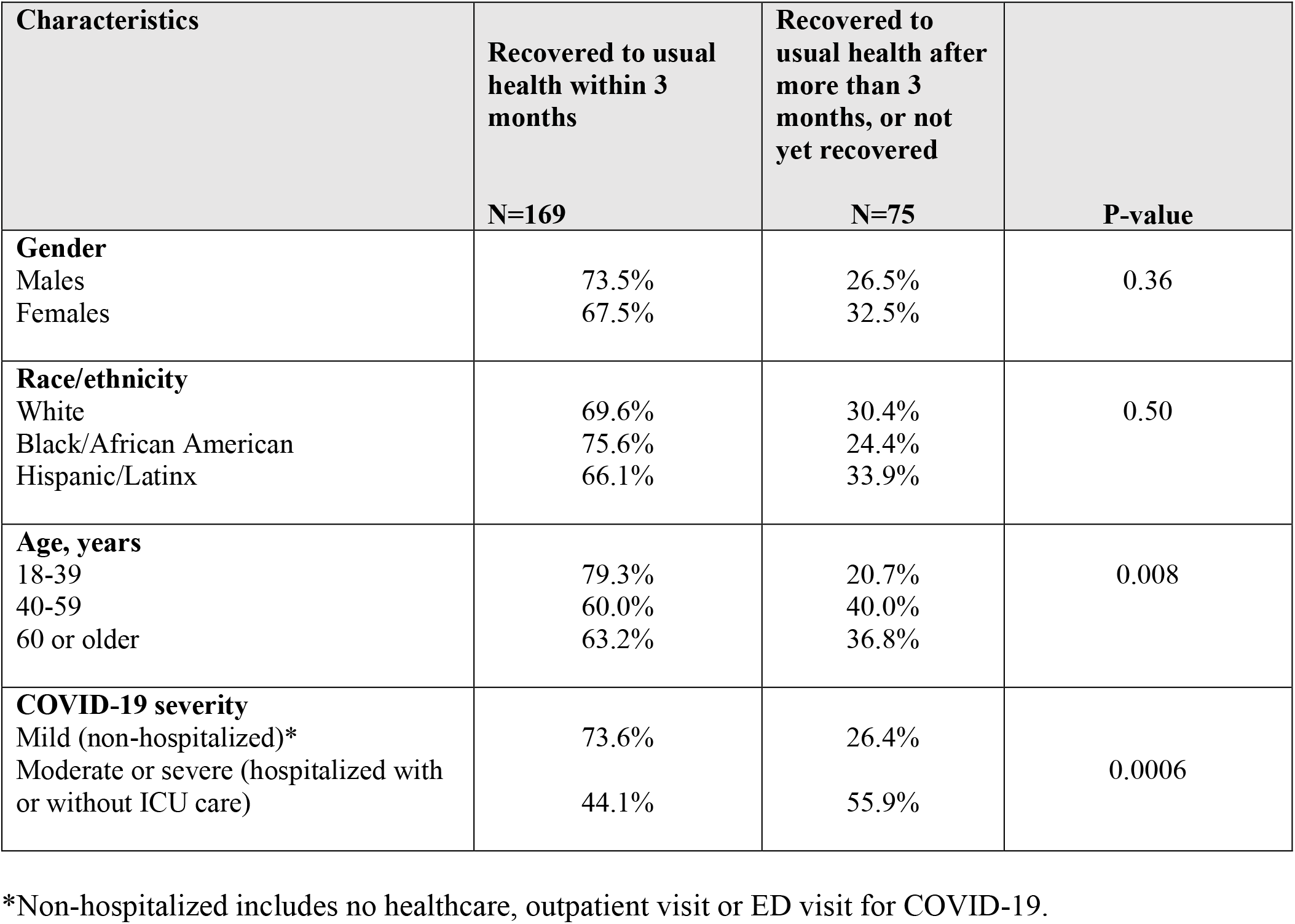
Recovery timeline, among participants >3 months since positive COVID-19 test

### Independent predictors of PASC-3 months

In a multivariable logistic regression model (Table 4), older age and greater severity of COVID-19 were independently associated with incomplete recovery. Participants who were 40-59 years were significantly less likely to return to usual health within 3 months (vs. age 18-39 years: adjusted odds ratio [aOR] 0.46, 95% confidence intervals [CI] 0.24 to 0.90). A similar pattern was noted in participants who were age 60 years and older vs. age 18-39 years, though differences were not significant. Participants who were hospitalized with COVID-19 were significantly less likely to return to usual health within 3 months (vs. not hospitalized: aOR 0.28, 95% CI 0.12 to 0.64). Gender, and race and ethnicity were not significantly associated with time to recovery in the multivariable regression model, though confidence intervals were wide suggesting additional data might provide evidence for these associations.

**Table 4:** Independent predictors of recovery to usual health within 3 months, multi-variable logistic regression models

| Characteristic | OR | 95% CI | p-value |
| --- | --- | --- | --- |
| **Gender** |  |  |  |
| Male (reference) | 1.0 (reference) | -- | -- |
| Female | 0.58 | 0.29-1.16 | 0.13 |
| **Race and ethnicity** |  |  |  |
| White (reference) | 1.0 (reference) | -- | -- |
| Black/African-American | 1.35 | 0.56-3.28 | 0.51 |
| Hispanic/Latinx | 0.77 | 0.39-1.53 | 0.45 |
| **Age** |  |  |  |
| 18-39 years | 1.0 (reference) |  |  |
| 40-59 years | 0.46 | 0.24-0.90 | 0.02 |
| 60 years or older | 0.58 | 0.24-1.42 | 0.24 |
| **Severity of COVID-19** |  |  |  |
| Mild (non-hospitalized) | 1.0 (reference) | -- | -- |
| Moderate or severe (hospitalized with or without ICU visit) | 0.28 | 0.12-0.64 | 0.003 |

OR= Odds ratio
95% CI = 95% confidence interval

## DISCUSSION

Our combination of community-engaged enrollment, COVID-19 research registries, and COVID-19 clinical registries helped to rapidly (over 7 days) recruit a predominantly minority (Hispanic/Latinx or Black/African-American) participant population, of whom about 85% had mild (no healthcare, outpatient care, or ED care), 9% had moderate (hospitalization without ICU-level care), and 5% had severe (hospitalization with ICU-level care) COVID-19. We found that about 1 in 4 participants did not return to their usual state of health by 3 months (PASC-3 months). Older age and more severe COVID-19 independently predicted PASC-3 months. We did not find significant gender, race or ethnic differences independently predicted the likelihood of PASC-3 months.

In the U.S., racial and ethnic minorities (e.g., Hispanic/Latinx and Black/African American populations) have a higher risk of SARS-CoV-2 infection, hospitalizations, and death.(10) Our approach to recruitment therefore used a combination of strategies, including collaborations with community partners, to rapidly recruit a diverse population including those at disproportionately high risk of morbidity and mortality from COVID-19. The community-engaged enrollment of participants has, to our knowledge, not yet been reported in other PASC studies. Such collaborations allowed us to enroll a sizable number of Hispanic/Latinx participants (N=188 [47% of n=401], with N=118 [48% of N=246] reporting a COVID-19 diagnosis 3 or more months prior to survey completion)--larger than in other US-based studies with Ns for Hispanic/Latinx participants below 100.(11-13, 18, 19) The community-engaged approach to recruitment also helped us to develop the infrastructure needed for broader dissemination of study findings (e.g., through virtual townhalls, in-person vaccination drives supported by community- and faith-based organizations) and for planning and implementing other stakeholder-supported studies.

Our multi-strategy approach to recruitment helped us to enroll participants across a spectrum (mild to severe) of acute illness associated with COVID-19, with about 85% of participants reporting mild COVID-19 (not hospitalized) and about 15% reporting moderate and severe COVID-19 (hospitalized with and without ICU-level care, respectively). The current report stands in contrast to most of the published literature, which has focused on post-hospital population (5-7, 19) or ambulatory population.(11-13, 20) Our findings about the patterns of PASC symptoms (e.g., fatigue, difficulty concentrating or thinking, lost of taste or smell) and increased risk of PASC among hospititalized patients are broadly consistent with previously published reports, suggesting that our multi-pronged approach to rapid recruitment did not result in widely divergent estimates.

There is no universally agreed-upon time frame after COVID-19 diagnosis for assessing symptoms of PASC; various studies have employed a wide range of time friames after COVID-19 diagnosis (e.g., 2 weeks to 6 months).(7-9, 11-13, 18-21) We selected a 3-month time frame to identify better the subset of individuals who may be most at risk for ongoing health concerns. This decision was based on concerns that a shorter time-frame (e.g., <1 month after COVID-19) may not adequately account for trajectories of recovery from acute illness regardless of cause, while a longer time-frame (e.g., 6 months or longer after COVID-19) would ignore multiple months of physical and mental health concerns. A recently published report found an increased risk of death and greater health care utilization after hospitalizations with COVID-19 persists for at least 6-months compared to hospitalizations with influenza.(11) Taken together with the pattern of some persistent symptoms (especially anosmia and dysgeusia > 3 months), these findings suggest that PASC is not simply another name for post-hospital syndrome.(22)

There were several strengths in our study design. The broad representation from community-based partners helped us to enroll within 7 days a diverse sample of patients with a history of COVID-19 that allows for a more time-sensitive, current snap-shot of the symptom and recovery experience of individuals with COVID-19. Given the dynamics of the changing landscape of the COVID-19 pandemic, vaccination rates, and emergence of new variants,(23-26) the ILLInet-PASC consortium appears is well-positioned to conduct rapid assessments of health, symptoms, and behavioral risk patterns that can be used to inform public health strategies and research priorities to mitigate the burden from COVID-19. Our partnerships with community- and faith-based organizations offer the potential to build on this work to recruit a larger sample of individuals to systematically examine race, ethnicity, comorbid illness, vaccination, and other potential determinants of recovery; such information can help to inform policymakers about the need for socioeconomic and health-related supports during the recovery phase after SARS-CoV-2 infection. We are also well-positioned to test the multi-pronged strategy of community-based enrollment, and clinical and research registries to enroll participants in high-dimensional prospective cohort studies that includes the collection of patient-reported information, biospecimens (e.g., blood, stool, saliva), physical and neurocognitive data using remote participant monitoring, and imaging data (e.g., brain, lungs, heart) to identify biologic targets for treatment and prevention studies.

Our study also has limitations. We relied on self-reported data, rather than laboratory-confirmed test results for SARS-CoV-2. Our study did not employ a population-based sampling strategy, so the prevalence and duration of different symptoms may not offer generalizable estimates across the state of Illinois. Moreover, our data were collected over a one week period in April 2021 and results may change with secular trends in the number of mild, moderate, and severe cases of COVID-19 cases. Also, the size of our one-week participant sample was insufficient to adequately examine gender, race and ethnicity as predictors of PASC at 3 months. A longer duration of survey collection or additional sources of participant populations will be needed to overcome limitations due to inadequate statistical power for some analyses.

In summary, we have demonstrated that the combination of community-engaged enrollment, COVID-19 research registries, and COVID-19 clinical registries can be used to rapidly recruit a predominantly minority participant population across the spectrum of COVID-19 severity. About 1 in 4 individuals had not recovered to their usual state of health after 3 months (even fewer among older individuals and those who had been hospitalized with COVID-19). Until there are proven-effective interventions to treat PASC, public health messaging should include the importance of preventing PASC through vaccination and other strategies to prevent infection with SARS-CoV-2. The partnerships with community- and faith-based organizations that were developed for the current study offer the potential to build on this work.

## Supporting information

COI disclosures

STROBE checklist

## Data Availability

All data referred to in the manuscript are available through the corresponding author

## DATA AVAILABLITY

All data referred to in the manuscript are available through the corresponding author.

## IRB REVIEW

The Institutional Review Boards at the University of Illinois Chicago (Protocol # 2021-0410) and Peoria (Protocol #1745454-1), Carle Hospital in Urbana (Protocol #21CRU3374), and Jesse Brown VA Medical Center (Protocol #162103-1) reviewed the ILLInet-PASC study protocol and determined the study to be exempt from on-going review because the data were de-identified at the time of collection.

