## Supplementary material for "Rapid implementation of cross-sectional study: Post-acute sequelae of SARS-CoV-2 (PASC) in a racially and ethnically diverse sample in Illinois": COI disclosures

### ICMJE DISCLOSURE FORM

**Date:** April 25, 2021
**Your Name:** Meenakshy Aiyer MD
**Manuscript Title:** Rapid implementation of cross-sectional study: Post-acute sequelae of SARS-CoV-2 (PASC) in an ethnically diverse sample in Illinois
**Manuscript number (if known):** Not known

**In the interest of transparency, we ask you to disclose all relationships/activities/interests listed below that are related to the content of your manuscript. "Related" means any relation with for-profit or not-for-profit third parties whose interests may be affected by the content of the manuscript. Disclosure represents a commitment to transparency and does not necessarily indicate a bias. If you are in doubt about whether to list a relationship/activity/interest, it is preferable that you do so.**

**The following questions apply to the author's relationships/activities/interests as they relate to the current manuscript only.**

**The author's relationships/activities/interests should be defined broadly. For example, if your manuscript pertains to the epidemiology of hypertension, you should declare all relationships with manufacturers of antihypertensive medication, even if that medication is not mentioned in the manuscript.**

**In item #1 below, report all support for the work reported in this manuscript without time limit. For all other items, the time frame for disclosure is the past 36 months.**

| | | Name all entities with whom you have this relationship or indicate none (add rows as needed) | Specifications/Comments (e.g., if payments were made to you or to your institution) |
|---|---|---|---|
| | | **Time frame: Since the initial planning of the work** | |
| 1 | All support for the present manuscript (e.g., funding, provision of study materials, medical writing, article processing charges, etc.) **No time limit for this item.** | ____None | |
| | | **Time frame: past 36 months** | |
| 2 | Grants or contracts from any entity (if not indicated in item #1 above). | | |
| 3 | Royalties or licenses | ____None | |
| 4 | Consulting fees | ____None | |

| | | |
|---|---|---|
| 5 | Payment or honoraria for lectures, presentations, speakers bureaus, manuscript writing or educational events | ____None |
| 6 | Payment for expert testimony | ____None |
| 7 | Support for attending meetings and/or travel | ____None |
| 8 | Patents planned, issued or pending | ____None |
| 9 | Participation on a Data Safety Monitoring Board or Advisory Board | ____None |
| 10 | Leadership or fiduciary role in other board, society, committee or advocacy group, paid or unpaid | Easter Seals Board Central Illinois |
| | | Community Foundation of Central Illinois |
| 11 | Stock or stock options | ____None |
| 12 | Receipt of equipment, materials, drugs, medical writing, gifts or other services | ____None |
| 13 | Other financial or non-financial interests | ____None |

**Please place an "X" next to the following statement to indicate your agreement:**

__X_ **I certify that I have answered every question and have not altered the wording of any of the questions on this form.**

### ICMJE DISCLOSURE FORM

**Date:** _4.27.21__
**Your Name:** __David Chestek__
**Manuscript Title:** Rapid implementation of cross-sectional study: Post-acute sequelae of SARS-CoV-2 (PASC) in an ethnically diverse sample in Illinois
**Manuscript number (if known):** Not known

**In the interest of transparency, we ask you to disclose all relationships/activities/interests listed below that are related to the content of your manuscript. "Related" means any relation with for-profit or not-for-profit third parties whose interests may be affected by the content of the manuscript. Disclosure represents a commitment to transparency and does not necessarily indicate a bias. If you are in doubt about whether to list a relationship/activity/interest, it is preferable that you do so.**

**The following questions apply to the author's relationships/activities/interests as they relate to the current manuscript only.**

**The author's relationships/activities/interests should be defined broadly. For example, if your manuscript pertains to the epidemiology of hypertension, you should declare all relationships with manufacturers of antihypertensive medication, even if that medication is not mentioned in the manuscript.**

**In item #1 below, report all support for the work reported in this manuscript without time limit. For all other items, the time frame for disclosure is the past 36 months.**

| | | **Name all entities with whom you have this relationship or indicate none (add rows as needed)** | **Specifications/Comments (e.g., if payments were made to you or to your institution)** |
|---|---|---|---|
| | | **Time frame: Since the initial planning of the work** | |
| 1 | All support for the present manuscript (e.g., funding, provision of study materials, medical writing, article processing charges, etc.) **No time limit for this item.** | None | |
| | | **Time frame: past 36 months** | |
| 2 | Grants or contracts from any entity (if not indicated in item #1 above). | NIH | |
| | | C3.ai | |
| | | CDC | National Heart, Lung and Blood Institute |
| 3 | Royalties or licenses | None | |
| 4 | Consulting fees | None | |

| | | |
|---|---|---|
| 5 | Payment or honoraria for lectures, presentations, speakers bureaus, manuscript writing or educational events | None |
| 6 | Payment for expert testimony | None |
| 7 | Support for attending meetings and/or travel | None |
| 8 | Patents planned, issued or pending | None |
| 9 | Participation on a Data Safety Monitoring Board or Advisory Board | None |
| 10 | Leadership or fiduciary role in other board, society, committee or advocacy group, paid or unpaid | None |
| 11 | Stock or stock options | None |
| 12 | Receipt of equipment, materials, drugs, medical writing, gifts or other services | None |
| 13 | Other financial or non-financial interests | None |

**Please place an "X" next to the following statement to indicate your agreement:**

**__X_ I certify that I have answered every question and have not altered the wording of any of the questions on this form.**

### ICMJE DISCLOSURE FORM

**Date: 4/26/21**
**Your Name: Christine Canfield**
**Manuscript Title:** Rapid implementation of cross-sectional study: Post-acute sequelae of SARS-CoV-2 (PASC) in an ethnically diverse sample in Illinois
**Manuscript number (if known):** Not known

**In the interest of transparency, we ask you to disclose all relationships/activities/interests listed below that are related to the content of your manuscript. "Related" means any relation with for-profit or not-for-profit third parties whose interests may be affected by the content of the manuscript. Disclosure represents a commitment to transparency and does not necessarily indicate a bias. If you are in doubt about whether to list a relationship/activity/interest, it is preferable that you do so.**

**The following questions apply to the author's relationships/activities/interests as they relate to the current manuscript only.**

**The author's relationships/activities/interests should be defined broadly. For example, if your manuscript pertains to the epidemiology of hypertension, you should declare all relationships with manufacturers of antihypertensive medication, even if that medication is not mentioned in the manuscript.**

**In item #1 below, report all support for the work reported in this manuscript without time limit. For all other items, the time frame for disclosure is the past 36 months.**

| | | Name all entities with whom you have this relationship or indicate none (add rows as needed) | Specifications/Comments (e.g., if payments were made to you or to your institution) |
|---|---|---|---|
| | | **Time frame: Since the initial planning of the work** | |
| 1 | All support for the present manuscript (e.g., funding, provision of study materials, medical writing, article processing charges, etc.) **No time limit for this item.** | X None | |
| | | **Time frame: past 36 months** | |
| 2 | Grants or contracts from any entity (if not indicated in item #1 above). | X None | |
| 3 | Royalties or licenses | X None | |
| 4 | Consulting fees | X None | |

| | | |
|---|---|---|
| 5 | Payment or honoraria for lectures, presentations, speakers bureaus, manuscript writing or educational events | X None |
| 6 | Payment for expert testimony | X None |
| 7 | Support for attending meetings and/or travel | X None |
| 8 | Patents planned, issued or pending | X None |
| 9 | Participation on a Data Safety Monitoring Board or Advisory Board | X None |
| 10 | Leadership or fiduciary role in other board, society, committee or advocacy group, paid or unpaid | X None |
| 11 | Stock or stock options | X None |
| 12 | Receipt of equipment, materials, drugs, medical writing, gifts or other services | X None |
| 13 | Other financial or non-financial interests | X None |

**Please place an "X" next to the following statement to indicate your agreement:**

**X I certify that I have answered every question and have not altered the wording of any of the questions on this form.**

#### ICMJE DISCLOSURE FORM

**Date: April 28, 2021**
**Your Name: Marina Del Rios**
**Manuscript Title:** Rapid implementation of cross-sectional study: Post-acute sequelae of SARS-CoV-2 (PASC) in an ethnically diverse sample in Illinois
**Manuscript number (if known):** Not known

**In the interest of transparency, we ask you to disclose all relationships/activities/interests listed below that are related to the content of your manuscript. "Related" means any relation with for-profit or not-for-profit third parties whose interests may be affected by the content of the manuscript. Disclosure represents a commitment to transparency and does not necessarily indicate a bias. If you are in doubt about whether to list a relationship/activity/interest, it is preferable that you do so.**

**The following questions apply to the author's relationships/activities/interests as they relate to the current manuscript only.**

**The author's relationships/activities/interests should be defined broadly. For example, if your manuscript pertains to the epidemiology of hypertension, you should declare all relationships with manufacturers of antihypertensive medication, even if that medication is not mentioned in the manuscript.**

**In item #1 below, report all support for the work reported in this manuscript without time limit. For all other items, the time frame for disclosure is the past 36 months.**

| | | **Name all entities with whom you have this relationship or indicate none (add rows as needed)** | **Specifications/Comments (e.g., if payments were made to you or to your institution)** |
|---|---|---|---|
| **Time frame: Since the initial planning of the work** | | | |
| 1 | All support for the present manuscript (e.g., funding, provision of study materials, medical writing, article processing charges, etc.) **No time limit for this item.** | ____None | |
| **Time frame: past 36 months** | | | |
| 2 | Grants or contracts from any entity (if not indicated in item #1 above). | ____None | |
| 3 | Royalties or licenses | ____None | |
| 4 | Consulting fees | ____None | |

| | | | |
|---|---|---|---|
| 5 | Payment or honoraria for lectures, presentations, speakers bureaus, manuscript writing or educational events | ____None | |
| 6 | Payment for expert testimony | ____None | |
| 7 | Support for attending meetings and/or travel | ____None | |
| 8 | Patents planned, issued or pending | ____None | |
| 9 | Participation on a Data Safety Monitoring Board or Advisory Board | ____None | |
| 10 | Leadership or fiduciary role in other board, society, committee or advocacy group, paid or unpaid | Illinois Unidos (Illinois Latino COVID19 Initiative) | Chair Health and Policy Committee, Unpaid Advocacy Group |
| 11 | Stock or stock options | ____None | |
| 12 | Receipt of equipment, materials, drugs, medical writing, gifts or other services | ____None | |
| 13 | Other financial or non-financial interests | ____None | |

**Please place an "X" next to the following statement to indicate your agreement:**

**_X_ I certify that I have answered every question and have not altered the wording of any of the questions on this form.**

### ICMJE DISCLOSURE FORM

**Date:** **_4/26/2021__**
**Your Name:** **_Judith A. Cook___**
**Manuscript Title:** Rapid implementation of cross-sectional study: Post-acute sequelae of SARS-CoV-2 (PASC) in an ethnically diverse sample in Illinois
**Manuscript number (if known):** Not known

**In the interest of transparency, we ask you to disclose all relationships/activities/interests listed below that are related to the content of your manuscript. "Related" means any relation with for-profit or not-for-profit third parties whose interests may be affected by the content of the manuscript. Disclosure represents a commitment to transparency and does not necessarily indicate a bias. If you are in doubt about whether to list a relationship/activity/interest, it is preferable that you do so.**

**The following questions apply to the author's relationships/activities/interests as they relate to the current manuscript only.**

**The author's relationships/activities/interests should be defined broadly. For example, if your manuscript pertains to the epidemiology of hypertension, you should declare all relationships with manufacturers of antihypertensive medication, even if that medication is not mentioned in the manuscript.**

**In item #1 below, report all support for the work reported in this manuscript without time limit. For all other items, the time frame for disclosure is the past 36 months.**

| | | Name all entities with whom you have this relationship or indicate none (add rows as needed) | Specifications/Comments (e.g., if payments were made to you or to your institution) |
|---|---|---|---|
| | | **Time frame: Since the initial planning of the work** | |
| 1 | All support for the present manuscript (e.g., funding, provision of study materials, medical writing, article processing charges, etc.) **No time limit for this item.** | ____None | |
| | | **Time frame: past 36 months** | |
| 2 | Grants or contracts from any entity (if not indicated in item #1 above). | ____None | |
| 3 | Royalties or licenses | ____None | |
| 4 | Consulting fees | ____None | |

| | | |
|---|---|---|
| 5 | Payment or honoraria for lectures, presentations, speakers bureaus, manuscript writing or educational events | ____None |
| 6 | Payment for expert testimony | ____None |
| 7 | Support for attending meetings and/or travel | ____None |
| 8 | Patents planned, issued or pending | ____None |
| 9 | Participation on a Data Safety Monitoring Board or Advisory Board | ____None |
| 10 | Leadership or fiduciary role in other board, society, committee or advocacy group, paid or unpaid | ____None |
| 11 | Stock or stock options | ____None |
| 12 | Receipt of equipment, materials, drugs, medical writing, gifts or other services | ____None |
| 13 | Other financial or non-financial interests | ____None |

**Please place an "X" next to the following statement to indicate your agreement:**

**_X__ I certify that I have answered every question and have not altered the wording of any of the questions on this form.**

#### ICMJE DISCLOSURE FORM

**Date:** April 25, 2021
**Your Name:** Kathleen R. Diviak, PhD
**Manuscript Title:** Rapid implementation of cross-sectional study: Post-acute sequelae of SARS-CoV-2 (PASC) in an ethnically diverse sample in Illinois
**Manuscript number (if known):** Not known

**In the interest of transparency, we ask you to disclose all relationships/activities/interests listed below that are related to the content of your manuscript. "Related" means any relation with for-profit or not-for-profit third parties whose interests may be affected by the content of the manuscript. Disclosure represents a commitment to transparency and does not necessarily indicate a bias. If you are in doubt about whether to list a relationship/activity/interest, it is preferable that you do so.**

**The following questions apply to the author's relationships/activities/interests as they relate to the current manuscript only.**

**The author's relationships/activities/interests should be defined broadly. For example, if your manuscript pertains to the epidemiology of hypertension, you should declare all relationships with manufacturers of antihypertensive medication, even if that medication is not mentioned in the manuscript.**

**In item #1 below, report all support for the work reported in this manuscript without time limit. For all other items, the time frame for disclosure is the past 36 months.**

| | | **Name all entities with whom you have this relationship or indicate none (add rows as needed)** | **Specifications/Comments (e.g., if payments were made to you or to your institution)** |
|---|---|---|---|
| | | **Time frame: Since the initial planning of the work** | |
| 1 | All support for the present manuscript (e.g., funding, provision of study materials, medical writing, article processing charges, etc.) **No time limit for this item.** | ____None | |
| | | **Time frame: past 36 months** | |
| 2 | Grants or contracts from any entity (if not indicated in item #1 above). | ____NIH | Institution |
| 3 | Royalties or licenses | ____None | |
| 4 | Consulting fees | ____None | |

| | | |
|---|---|---|
| 5 | Payment or honoraria for lectures, presentations, speakers bureaus, manuscript writing or educational events | ____None |
| 6 | Payment for expert testimony | ____None |
| 7 | Support for attending meetings and/or travel | ____None |
| 8 | Patents planned, issued or pending | ____None |
| 9 | Participation on a Data Safety Monitoring Board or Advisory Board | ____None |
| 10 | Leadership or fiduciary role in other board, society, committee or advocacy group, paid or unpaid | ____None |
| 11 | Stock or stock options | ____None |
| 12 | Receipt of equipment, materials, drugs, medical writing, gifts or other services | ____None |
| 13 | Other financial or non-financial interests | ____None |

**Please place an "X" next to the following statement to indicate your agreement:**

**_X__ I certify that I have answered every question and have not altered the wording of any of the questions on this form.**

#### ICMJE DISCLOSURE FORM

**Date:** **April 27, 2021___**
**Your Name:** **_Angela M Ellison___**
**Manuscript Title:** Rapid implementation of cross-sectional study: Post-acute sequelae of SARS-CoV-2 (PASC) in an ethnically diverse sample in Illinois
**Manuscript number (if known):** Not known

**In the interest of transparency, we ask you to disclose all relationships/activities/interests listed below that are related to the content of your manuscript. "Related" means any relation with for-profit or not-for-profit third parties whose interests may be affected by the content of the manuscript. Disclosure represents a commitment to transparency and does not necessarily indicate a bias. If you are in doubt about whether to list a relationship/activity/interest, it is preferable that you do so.**

**The following questions apply to the author's relationships/activities/interests as they relate to the current manuscript only.**

**The author's relationships/activities/interests should be defined broadly. For example, if your manuscript pertains to the epidemiology of hypertension, you should declare all relationships with manufacturers of antihypertensive medication, even if that medication is not mentioned in the manuscript.**

**In item #1 below, report all support for the work reported in this manuscript without time limit. For all other items, the time frame for disclosure is the past 36 months.**

| | | **Name all entities with whom you have this relationship or indicate none (add rows as needed)** | **Specifications/Comments (e.g., if payments were made to you or to your institution)** |
|---|---|---|---|
| | | **Time frame: Since the initial planning of the work** | |
| 1 | All support for the present manuscript (e.g., funding, provision of study materials, medical writing, article processing charges, etc.) **No time limit for this item.** | ____None | |
| | | **Time frame: past 36 months** | |
| 2 | Grants or contracts from any entity (if not indicated in item #1 above). | ____None | |
| 3 | Royalties or licenses | ____None | |
| 4 | Consulting fees | ____None | |

| | | |
|---|---|---|
| 5 | Payment or honoraria for lectures, presentations, speakers bureaus, manuscript writing or educational events | ____None |
| 6 | Payment for expert testimony | ____None |
| 7 | Support for attending meetings and/or travel | ____None |
| 8 | Patents planned, issued or pending | ____None |
| 9 | Participation on a Data Safety Monitoring Board or Advisory Board | ____None |
| 10 | Leadership or fiduciary role in other board, society, committee or advocacy group, paid or unpaid | Illinois Community Heath Worker Association |
| 11 | Stock or stock options | ____None |
| 12 | Receipt of equipment, materials, drugs, medical writing, gifts or other services | ____None |
| 13 | Other financial or non-financial interests | ____None |

**Please place an "X" next to the following statement to indicate your agreement:**

**__X_ I certify that I have answered every question and have not altered the wording of any of the questions on this form.**

### ICMJE DISCLOSURE FORM

**Date: ___4-27-2021**
**Your Name: ____Howard S Gordon, MD**
**Manuscript Title:** Rapid implementation of cross-sectional study: Post-acute sequelae of SARS-CoV-2 (PASC) in an ethnically diverse sample in Illinois
**Manuscript number (if known):** Not known

**In the interest of transparency, we ask you to disclose all relationships/activities/interests listed below that are related to the content of your manuscript. "Related" means any relation with for-profit or not-for-profit third parties whose interests may be affected by the content of the manuscript. Disclosure represents a commitment to transparency and does not necessarily indicate a bias. If you are in doubt about whether to list a relationship/activity/interest, it is preferable that you do so.**

**The following questions apply to the author's relationships/activities/interests as they relate to the current manuscript only.**

**The author's relationships/activities/interests should be defined broadly. For example, if your manuscript pertains to the epidemiology of hypertension, you should declare all relationships with manufacturers of antihypertensive medication, even if that medication is not mentioned in the manuscript.**

**In item #1 below, report all support for the work reported in this manuscript without time limit. For all other items, the time frame for disclosure is the past 36 months.**

| | | **Name all entities with whom you have this relationship or indicate none (add rows as needed)** | **Specifications/Comments (e.g., if payments were made to you or to your institution)** |
|---|---|---|---|
| | | **Time frame: Since the initial planning of the work** | |
| 1 | All support for the present manuscript (e.g., funding, provision of study materials, medical writing, article processing charges, etc.) **No time limit for this item.** | ____None | |
| | | Jesse Brown VA Medical Center, Chicago, IL, USA | This material is the result of work supported in part with resources and the use of facilities at the Jesse Brown VA Medical Center, Chicago, Illinois. |
| | | Health Services Research and Development, Office of Research and Development, US Dept. of Veterans Affairs | This study was supported using data from the VA COVID-19 Shared Data Resource |
| | | **Time frame: past 36 months** | |
| 2 | | ____None | |

| | | |
|---|---|---|
| | Grants or contracts from any entity (if not indicated in item #1 above). | VA Health Services Research and Development |
| | | Patient Centered Outcomes Research Institute |
| | | National Institute of Health |
| 3 | Royalties or licenses | __x__None |
| 4 | Consulting fees | __x__None |
| 5 | Payment or honoraria for lectures, presentations, speakers bureaus, manuscript writing or educational events | __x__None |
| 6 | Payment for expert testimony | __x__None |
| 7 | Support for attending meetings and/or travel | __x__None |
| 8 | Patents planned, issued or pending | __x__None |
| 9 | Participation on a Data Safety Monitoring Board or Advisory Board | __x__None |
| 10 | Leadership or fiduciary role in other board, society, committee or advocacy group, paid or unpaid | __x__None |
| 11 | Stock or stock options | __x__None |
| 12 | Receipt of equipment, materials, drugs, medical writing, gifts or other services | __x__None |
| 13 | Other financial or non-financial interests | __x__None |

**Please place an "X" next to the following statement to indicate your agreement:**

**_x__ I certify that I have answered every question and have not altered the wording of any of the questions on this**

**form.**

#### ICMJE DISCLOSURE FORM

**Date: 4/27/2021**
**Your Name: Sai Dheeraj Illendula**
**Manuscript Title:** Rapid implementation of cross-sectional study: Post-acute sequelae of SARS-CoV-2 (PASC) in an ethnically diverse sample in Illinois
**Manuscript number (if known):** Not known

**In the interest of transparency, we ask you to disclose all relationships/activities/interests listed below that are related to the content of your manuscript. "Related" means any relation with for-profit or not-for-profit third parties whose interests may be affected by the content of the manuscript. Disclosure represents a commitment to transparency and does not necessarily indicate a bias. If you are in doubt about whether to list a relationship/activity/interest, it is preferable that you do so.**

**The following questions apply to the author's relationships/activities/interests as they relate to the current manuscript only.**

**The author's relationships/activities/interests should be defined broadly. For example, if your manuscript pertains to the epidemiology of hypertension, you should declare all relationships with manufacturers of antihypertensive medication, even if that medication is not mentioned in the manuscript.**

**In item #1 below, report all support for the work reported in this manuscript without time limit. For all other items, the time frame for disclosure is the past 36 months.**

| | | **Name all entities with whom you have this relationship or indicate none (add rows as needed)** | **Specifications/Comments (e.g., if payments were made to you or to your institution)** |
|---|---|---|---|
| | | **Time frame: Since the initial planning of the work** | |
| 1 | All support for the present manuscript (e.g., funding, provision of study materials, medical writing, article processing charges, etc.) **No time limit for this item.** | ____None | |
| | | **Time frame: past 36 months** | |
| 2 | Grants or contracts from any entity (if not indicated in item #1 above). | ____None | |
| 3 | Royalties or licenses | ____None | |
| 4 | Consulting fees | ____None | |

| | | |
|---|---|---|
| 5 | Payment or honoraria for lectures, presentations, speakers bureaus, manuscript writing or educational events | ____None |
| 6 | Payment for expert testimony | ____None |
| 7 | Support for attending meetings and/or travel | ____None |
| 8 | Patents planned, issued or pending | ____None |
| 9 | Participation on a Data Safety Monitoring Board or Advisory Board | ____None |
| 10 | Leadership or fiduciary role in other board, society, committee or advocacy group, paid or unpaid | ____None |
| 11 | Stock or stock options | ____None |
| 12 | Receipt of equipment, materials, drugs, medical writing, gifts or other services | ____None |
| 13 | Other financial or non-financial interests | ____None |

**Please place an "X" next to the following statement to indicate your agreement:**

**Sai Dheeraj Illendula, I certify that I have answered every question and have not altered the wording of any of the questions on this form.**

### ICMJE DISCLOSURE FORM

**Date:** **_April 28 2021__**
**Your Name: Manasa Kandula, MD**
**Manuscript Title:** Rapid implementation of cross-sectional study: Post-acute sequelae of SARS-CoV-2 (PASC) in an ethnically diverse sample in Illinois
**Manuscript number (if known):** Not known

**In the interest of transparency, we ask you to disclose all relationships/activities/interests listed below that are related to the content of your manuscript. "Related" means any relation with for-profit or not-for-profit third parties whose interests may be affected by the content of the manuscript. Disclosure represents a commitment to transparency and does not necessarily indicate a bias. If you are in doubt about whether to list a relationship/activity/interest, it is preferable that you do so.**

**The following questions apply to the author's relationships/activities/interests as they relate to the current manuscript only.**

**The author's relationships/activities/interests should be defined broadly. For example, if your manuscript pertains to the epidemiology of hypertension, you should declare all relationships with manufacturers of antihypertensive medication, even if that medication is not mentioned in the manuscript.**

**In item #1 below, report all support for the work reported in this manuscript without time limit. For all other items, the time frame for disclosure is the past 36 months.**

| | | Name all entities with whom you have this relationship or indicate none (add rows as needed) | Specifications/Comments (e.g., if payments were made to you or to your institution) |
|---|---|---|---|
| | | **Time frame: Since the initial planning of the work** | |
| 1 | All support for the present manuscript (e.g., funding, provision of study materials, medical writing, article processing charges, etc.) **No time limit for this item.** | ____None | |
| | | **Time frame: past 36 months** | |
| 2 | Grants or contracts from any entity (if not indicated in item #1 above). | ____None | |
| 3 | Royalties or licenses | ____None | |
| 4 | Consulting fees | ____None | |

| | | |
|---|---|---|
| 5 | Payment or honoraria for lectures, presentations, speakers bureaus, manuscript writing or educational events | ____None |
| 6 | Payment for expert testimony | ____None |
| 7 | Support for attending meetings and/or travel | ____None |
| 8 | Patents planned, issued or pending | ____None |
| 9 | Participation on a Data Safety Monitoring Board or Advisory Board | ____None |
| 10 | Leadership or fiduciary role in other board, society, committee or advocacy group, paid or unpaid | ____None |
| 11 | Stock or stock options | ____None |
| 12 | Receipt of equipment, materials, drugs, medical writing, gifts or other services | ____None |
| 13 | Other financial or non-financial interests | ____None |

**Please place an "X" next to the following statement to indicate your agreement:**

**__X_ I certify that I have answered every question and have not altered the wording of any of the questions on this form.**

### ICMJE DISCLOSURE FORM

**Date:** **_April 28, 2021__**
**Your Name:** **___Jonathan D. Klein_**
**Manuscript Title:** Rapid implementation of cross-sectional study: Post-acute sequelae of SARS-CoV-2 (PASC) in an ethnically diverse sample in Illinois
**Manuscript number (if known):** Not known

**In the interest of transparency, we ask you to disclose all relationships/activities/interests listed below that are related to the content of your manuscript. "Related" means any relation with for-profit or not-for-profit third parties whose interests may be affected by the content of the manuscript. Disclosure represents a commitment to transparency and does not necessarily indicate a bias. If you are in doubt about whether to list a relationship/activity/interest, it is preferable that you do so.**

**The following questions apply to the author's relationships/activities/interests as they relate to the current manuscript only.**

**The author's relationships/activities/interests should be defined broadly. For example, if your manuscript pertains to the epidemiology of hypertension, you should declare all relationships with manufacturers of antihypertensive medication, even if that medication is not mentioned in the manuscript.**

**In item #1 below, report all support for the work reported in this manuscript without time limit. For all other items, the time frame for disclosure is the past 36 months.**

| | | **Name all entities with whom you have this relationship or indicate none (add rows as needed)** | **Specifications/Comments (e.g., if payments were made to you or to your institution)** |
|---|---|---|---|
| | | **Time frame: Since the initial planning of the work** | |
| 1 | All support for the present manuscript (e.g., funding, provision of study materials, medical writing, article processing charges, etc.) **No time limit for this item.** | ____None | |
| | | **Time frame: past 36 months** | |
| 2 | Grants or contracts from any entity (if not indicated in item #1 above). | ____None | |
| 3 | Royalties or licenses | ____None | |
| 4 | Consulting fees | ____None | |

| | | |
|---|---|---|
| 5 | Payment or honoraria for lectures, presentations, speakers bureaus, manuscript writing or educational events | ____None |
| 6 | Payment for expert testimony | ____None |
| 7 | Support for attending meetings and/or travel | ____None |
| 8 | Patents planned, issued or pending | ____None |
| 9 | Participation on a Data Safety Monitoring Board or Advisory Board | ____None |
| 10 | Leadership or fiduciary role in other board, society, committee or advocacy group, paid or unpaid | ____None |
| 11 | Stock or stock options | ____None |
| 12 | Receipt of equipment, materials, drugs, medical writing, gifts or other services | ____None |
| 13 | Other financial or non-financial interests | ____None |

**Please place an "X" next to the following statement to indicate your agreement:**

**_xx__ I certify that I have answered every question and have not altered the wording of any of the questions on this form.**

### ICMJE DISCLOSURE FORM

**Date:** April 25, 2021
**Your Name:** Jerry A. Krishnan, MD, PhD
**Manuscript Title:** Rapid implementation of cross-sectional study: Post-acute sequelae of SARS-CoV-2 (PASC) in an ethnically diverse sample in Illinois
**Manuscript number (if known):** Not known

**In the interest of transparency, we ask you to disclose all relationships/activities/interests listed below that are related to the content of your manuscript. "Related" means any relation with for-profit or not-for-profit third parties whose interests may be affected by the content of the manuscript. Disclosure represents a commitment to transparency and does not necessarily indicate a bias. If you are in doubt about whether to list a relationship/activity/interest, it is preferable that you do so.**

**The following questions apply to the author's relationships/activities/interests as they relate to the current manuscript only.**

**The author's relationships/activities/interests should be defined broadly. For example, if your manuscript pertains to the epidemiology of hypertension, you should declare all relationships with manufacturers of antihypertensive medication, even if that medication is not mentioned in the manuscript.**

**In item #1 below, report all support for the work reported in this manuscript without time limit. For all other items, the time frame for disclosure is the past 36 months.**

| | | **Name all entities with whom you have this relationship or indicate none (add rows as needed)** | **Specifications/Comments (e.g., if payments were made to you or to your institution)** |
|---|---|---|---|
| | | **Time frame: Since the initial planning of the work** | |
| 1 | All support for the present manuscript (e.g., funding, provision of study materials, medical writing, article processing charges, etc.) **No time limit for this item.** | ____None | |
| | | **Time frame: past 36 months** | |
| 2 | Grants or contracts from any entity (if not indicated in item #1 above). | ____NIH | Institution |
| | | Sergey Brin Family Foundation | Institution |
| | | Patient Centered Outcomes Research Institutee | Institution |
| 3 | Royalties or licenses | ____None | |

| | | | |
|---|---|---|---|
| 4 | Consulting fees | ____None | |
| 5 | Payment or honoraria for lectures, presentations, speakers bureaus, manuscript writing or educational events | ____None | |
| 6 | Payment for expert testimony | ____None | |
| 7 | Support for attending meetings and/or travel | ____None | |
| 8 | Patents planned, issued or pending | ____None | |
| 9 | Participation on a Data Safety Monitoring Board or Advisory Board | ____None | |
| 10 | Leadership or fiduciary role in other board, society, committee or advocacy group, paid or unpaid | Respiratory Health Association | Unpaid |
| | | COPD Foundation | Unpaid |
| 11 | Stock or stock options | ____None | |
| 12 | Receipt of equipment, materials, drugs, medical writing, gifts or other services | ____None | |
| 13 | Other financial or non-financial interests | ____None | |

**Please place an "X" next to the following statement to indicate your agreement:**

**__X_ I certify that I have answered every question and have not altered the wording of any of the questions on this form.**

### ICMJE DISCLOSURE FORM

**Date:** _4.26.21__
**Your Name:** Karen Larimer____
**Manuscript Title:** Rapid implementation of cross-sectional study: Post-acute sequelae of SARS-CoV-2 (PASC) in an ethnically diverse sample in Illinois
**Manuscript number (if known):** Not known

**In the interest of transparency, we ask you to disclose all relationships/activities/interests listed below that are related to the content of your manuscript. "Related" means any relation with for-profit or not-for-profit third parties whose interests may be affected by the content of the manuscript. Disclosure represents a commitment to transparency and does not necessarily indicate a bias. If you are in doubt about whether to list a relationship/activity/interest, it is preferable that you do so.**

**The following questions apply to the author's relationships/activities/interests as they relate to the current manuscript only.**

**The author's relationships/activities/interests should be defined broadly. For example, if your manuscript pertains to the epidemiology of hypertension, you should declare all relationships with manufacturers of antihypertensive medication, even if that medication is not mentioned in the manuscript.**

**In item #1 below, report all support for the work reported in this manuscript without time limit. For all other items, the time frame for disclosure is the past 36 months.**

| | | **Name all entities with whom you have this relationship or indicate none (add rows as needed)** | **Specifications/Comments (e.g., if payments were made to you or to your institution)** |
|---|---|---|---|
| | | **Time frame: Since the initial planning of the work** | |
| 1 | All support for the present manuscript (e.g., funding, provision of study materials, medical writing, article processing charges, etc.) **No time limit for this item.** | none | |
| | | **Time frame: past 36 months** | |
| 2 | Grants or contracts from any entity (if not indicated in item #1 above). | physIQ | Employee and equity holder of physIQ |
| | | NIH | DeCODe Study CONTRACT No. 75N91020C00040 |
| 3 | Royalties or licenses | ____None | |
| 4 | Consulting fees | ____None | |

| | | |
|---|---|---|
| 5 | Payment or honoraria for lectures, presentations, speakers bureaus, manuscript writing or educational events | ____None |
| 6 | Payment for expert testimony | ____None |
| 7 | Support for attending meetings and/or travel | ____None |
| 8 | Patents planned, issued or pending | ____None |
| 9 | Participation on a Data Safety Monitoring Board or Advisory Board | ____None |
| 10 | Leadership or fiduciary role in other board, society, committee or advocacy group, paid or unpaid | American Heart Association |
| | | Preventive Cardiovascular Nurse Association |
| 11 | Stock or stock options | ____None |
| 12 | Receipt of equipment, materials, drugs, medical writing, gifts or other services | ____None |
| 13 | Other financial or non-financial interests | ____None |

**Please place an "X" next to the following statement to indicate your agreement:**

**_X__ I certify that I have answered every question and have not altered the wording of any of the questions on this form.**

### ICMJE DISCLOSURE FORM

**Date:** April 25, 2021
**Your Name:** James Lash, MD
**Manuscript Title:** Rapid implementation of cross-sectional study: Post-acute sequelae of SARS-CoV-2 (PASC) in an ethnically diverse sample in Illinois
**Manuscript number (if known):** Not known

**In the interest of transparency, we ask you to disclose all relationships/activities/interests listed below that are related to the content of your manuscript. "Related" means any relation with for-profit or not-for-profit third parties whose interests may be affected by the content of the manuscript. Disclosure represents a commitment to transparency and does not necessarily indicate a bias. If you are in doubt about whether to list a relationship/activity/interest, it is preferable that you do so.**

**The following questions apply to the author's relationships/activities/interests as they relate to the current manuscript only.**

**The author's relationships/activities/interests should be defined broadly. For example, if your manuscript pertains to the epidemiology of hypertension, you should declare all relationships with manufacturers of antihypertensive medication, even if that medication is not mentioned in the manuscript.**

**In item #1 below, report all support for the work reported in this manuscript without time limit. For all other items, the time frame for disclosure is the past 36 months.**

| | | Name all entities with whom you have this relationship or indicate none (add rows as needed) | Specifications/Comments (e.g., if payments were made to you or to your institution) |
|---|---|---|---|
| **Time frame: Since the initial planning of the work** | | | |
| 1 | All support for the present manuscript (e.g., funding, provision of study materials, medical writing, article processing charges, etc.) **No time limit for this item.** | ____None | |
| **Time frame: past 36 months** | | | |
| 2 | Grants or contracts from any entity (if not indicated in item #1 above). | ____NIH | Institution |
| 3 | Royalties or licenses | ____None | |
| 4 | Consulting fees | ____None | |

| | | |
|---|---|---|
| 5 | Payment or honoraria for lectures, presentations, speakers bureaus, manuscript writing or educational events | ____None |
| 6 | Payment for expert testimony | ____None |
| 7 | Support for attending meetings and/or travel | ____None |
| 8 | Patents planned, issued or pending | ____None |
| 9 | Participation on a Data Safety Monitoring Board or Advisory Board | ____None |
| 10 | Leadership or fiduciary role in other board, society, committee or advocacy group, paid or unpaid | |
| 11 | Stock or stock options | ____None |
| 12 | Receipt of equipment, materials, drugs, medical writing, gifts or other services | ____None |
| 13 | Other financial or non-financial interests | ____None |

**Please place an "X" next to the following statement to indicate your agreement:**

__X_ **I certify that I have answered every question and have not altered the wording of any of the questions on this form.**

### ICMJE DISCLOSURE FORM

**Date:** 4/26/2021
**Your Name:** Janet Lin
**Manuscript Title:** Rapid implementation of cross-sectional study: Post-acute sequelae of SARS-CoV-2 (PASC) in an ethnically diverse sample in Illinois
**Manuscript number (if known):** Not known

**In the interest of transparency, we ask you to disclose all relationships/activities/interests listed below that are related to the content of your manuscript. "Related" means any relation with for-profit or not-for-profit third parties whose interests may be affected by the content of the manuscript. Disclosure represents a commitment to transparency and does not necessarily indicate a bias. If you are in doubt about whether to list a relationship/activity/interest, it is preferable that you do so.**

**The following questions apply to the author's relationships/activities/interests as they relate to the current manuscript only.**

**The author's relationships/activities/interests should be defined broadly. For example, if your manuscript pertains to the epidemiology of hypertension, you should declare all relationships with manufacturers of antihypertensive medication, even if that medication is not mentioned in the manuscript.**

**In item #1 below, report all support for the work reported in this manuscript without time limit. For all other items, the time frame for disclosure is the past 36 months.**

| | | Name all entities with whom you have this relationship or indicate none (add rows as needed) | Specifications/Comments (e.g., if payments were made to you or to your institution) |
|---|---|---|---|
| | | **Time frame: Since the initial planning of the work** | |
| 1 | All support for the present manuscript (e.g., funding, provision of study materials, medical writing, article processing charges, etc.) **No time limit for this item.** | ____None | |
| | | **Time frame: past 36 months** | |
| 2 | Grants or contracts from any entity (if not indicated in item #1 above). | NIH | Institution |
| | | Sergey Brin Family Foundation | Institution |
| 3 | Royalties or licenses | ____None | |
| 4 | Consulting fees | ____None | |

| | | | |
|---|---|---|---|
| 5 | Payment or honoraria for lectures, presentations, speakers bureaus, manuscript writing or educational events | ____None | |
| 6 | Payment for expert testimony | ____None | |
| 7 | Support for attending meetings and/or travel | ____None | |
| 8 | Patents planned, issued or pending | ____None | |
| 9 | Participation on a Data Safety Monitoring Board or Advisory Board | ____None | |
| 10 | Leadership or fiduciary role in other board, society, committee or advocacy group, paid or unpaid | Illinois College of Emergency Physicians | Board member, unpaid |
| | | AIDS Foundation of Chicago | Board member, unpaid |
| | | Chicago Board of Health | Board member, unpaid |
| 11 | Stock or stock options | ____None | |
| 12 | Receipt of equipment, materials, drugs, medical writing, gifts or other services | ____None | |
| 13 | Other financial or non-financial interests | ____None | |

**Please place an "X" next to the following statement to indicate your agreement:**

**__x_ I certify that I have answered every question and have not altered the wording of any of the questions on this form.**

### ICMJE DISCLOSURE FORM

**Date:** _4/26/21__
**Your Name:** __Jeffrey A. Loeb, MD PhD
**Manuscript Title:** Rapid implementation of cross-sectional study: Post-acute sequelae of SARS-CoV-2 (PASC) in an ethnically diverse sample in Illinois
**Manuscript number (if known):** Not known

**In the interest of transparency, we ask you to disclose all relationships/activities/interests listed below that are related to the content of your manuscript. "Related" means any relation with for-profit or not-for-profit third parties whose interests may be affected by the content of the manuscript. Disclosure represents a commitment to transparency and does not necessarily indicate a bias. If you are in doubt about whether to list a relationship/activity/interest, it is preferable that you do so.**

**The following questions apply to the author's relationships/activities/interests as they relate to the current manuscript only.**

**The author's relationships/activities/interests should be defined broadly. For example, if your manuscript pertains to the epidemiology of hypertension, you should declare all relationships with manufacturers of antihypertensive medication, even if that medication is not mentioned in the manuscript.**

**In item #1 below, report all support for the work reported in this manuscript without time limit. For all other items, the time frame for disclosure is the past 36 months.**

| | | Name all entities with whom you have this relationship or indicate none (add rows as needed) | Specifications/Comments (e.g., if payments were made to you or to your institution) |
|---|---|---|---|
| | | **Time frame: Since the initial planning of the work** | |
| 1 | All support for the present manuscript (e.g., funding, provision of study materials, medical writing, article processing charges, etc.) **No time limit for this item.** | ____None | |
| | | **Time frame: past 36 months** | |
| 2 | Grants or contracts from any entity (if not indicated in item #1 above). | NIH | Institution |
| | | DOD/CURE | Institution |
| | | MTPharma/Falk/CBC/DPI | Institution |
| 3 | Royalties or licenses | ____None | |
| 4 | Consulting fees | ____None | |

| | | | |
|---|---|---|---|
| 5 | Payment or honoraria for lectures, presentations, speakers bureaus, manuscript writing or educational events | ____None | |
| 6 | Payment for expert testimony | ____None | |
| 7 | Support for attending meetings and/or travel | ____None | |
| 8 | Patents planned, issued or pending | | Loeb JA and Ellis Kirchner A, “Method of Treating Epilepsy: DUSP4 activation with new use of formoterol to prevent and treat epilepsy,” USP Filed 5/20 UIC 202-142-01. |
| | | | Loeb JA and Song F “METHOD OF TREATING NEUORDEGENERATIVE DISEASE,” 1/10/19 application 62787852. |
| 9 | Participation on a Data Safety Monitoring Board or Advisory Board | ____None | |
| 10 | Leadership or fiduciary role in other board, society, committee or advocacy group, paid or unpaid | Chief Clinical Strategist | The Sturge Weber Foundation |
| 11 | Stock or stock options | ____None | |
| 12 | Receipt of equipment, materials, drugs, medical writing, gifts or other services | ____None | |
| 13 | Other financial or non-financial interests | ____None | |

**Please place an “X” next to the following statement to indicate your agreement:**

**_X__ I certify that I have answered every question and have not altered the wording of any of the questions on this form.**

#### ICMJE DISCLOSURE FORM

**Date:** April 28, 2021
**Your Name:** Teresa J. Lynch, MD
**Manuscript Title:** Rapid implementation of cross-sectional study: Post-acute sequelae of SARS-CoV-2 (PASC) in an ethnically diverse sample in Illinois
**Manuscript number (if known):** Not known

**In the interest of transparency, we ask you to disclose all relationships/activities/interests listed below that are related to the content of your manuscript. "Related" means any relation with for-profit or not-for-profit third parties whose interests may be affected by the content of the manuscript. Disclosure represents a commitment to transparency and does not necessarily indicate a bias. If you are in doubt about whether to list a relationship/activity/interest, it is preferable that you do so.**

**The following questions apply to the author's relationships/activities/interests as they relate to the current manuscript only.**

**The author's relationships/activities/interests should be defined broadly. For example, if your manuscript pertains to the epidemiology of hypertension, you should declare all relationships with manufacturers of antihypertensive medication, even if that medication is not mentioned in the manuscript.**

**In item #1 below, report all support for the work reported in this manuscript without time limit. For all other items, the time frame for disclosure is the past 36 months.**

| | | **Name all entities with whom you have this relationship or indicate none (add rows as needed)** | **Specifications/Comments (e.g., if payments were made to you or to your institution)** |
|---|---|---|---|
| | | **Time frame: Since the initial planning of the work** | |
| 1 | All support for the present manuscript (e.g., funding, provision of study materials, medical writing, article processing charges, etc.) **No time limit for this item.** | ____None | |
| | | **Time frame: past 36 months** | |
| 2 | Grants or contracts from any entity (if not indicated in item #1 above). | ____None | |
| 3 | Royalties or licenses | ____None | |
| 4 | Consulting fees | ____None | |

| | | |
|---|---|---|
| 5 | Payment or honoraria for lectures, presentations, speakers bureaus, manuscript writing or educational events | ____None |
| 6 | Payment for expert testimony | ____None |
| 7 | Support for attending meetings and/or travel | ____None |
| 8 | Patents planned, issued or pending | ____None |
| 9 | Participation on a Data Safety Monitoring Board or Advisory Board | ____None |
| 10 | Leadership or fiduciary role in other board, society, committee or advocacy group, paid or unpaid | ____None |
| 11 | Stock or stock options | ____None |
| 12 | Receipt of equipment, materials, drugs, medical writing, gifts or other services | ____None |
| 13 | Other financial or non-financial interests | ____None |

**Please place an "X" next to the following statement to indicate your agreement:**

**_X_ I certify that I have answered every question and have not altered the wording of any of the questions on this form.**

#### ICMJE DISCLOSURE FORM

**Date: __April 26, 2021_**
**Your Name: ___Robin Mermelstein, Ph.D._**
**Manuscript Title: Rapid implementation of cross-sectional study: Post-acute sequelae of SARS-CoV-2 (PASC) in an ethnically diverse sample in Illinois**
**Manuscript number (if known):** Not known

**In the interest of transparency, we ask you to disclose all relationships/activities/interests listed below that are related to the content of your manuscript. "Related" means any relation with for-profit or not-for-profit third parties whose interests may be affected by the content of the manuscript. Disclosure represents a commitment to transparency and does not necessarily indicate a bias. If you are in doubt about whether to list a relationship/activity/interest, it is preferable that you do so.**

**The following questions apply to the author's relationships/activities/interests as they relate to the current manuscript only.**

**The author's relationships/activities/interests should be defined broadly. For example, if your manuscript pertains to the epidemiology of hypertension, you should declare all relationships with manufacturers of antihypertensive medication, even if that medication is not mentioned in the manuscript.**

**In item #1 below, report all support for the work reported in this manuscript without time limit. For all other items, the time frame for disclosure is the past 36 months.**

| | | **Name all entities with whom you have this relationship or indicate none (add rows as needed)** | **Specifications/Comments (e.g., if payments were made to you or to your institution)** |
|---|---|---|---|
| | | **Time frame: Since the initial planning of the work** | |
| 1 | All support for the present manuscript (e.g., funding, provision of study materials, medical writing, article processing charges, etc.) **No time limit for this item.** | ___NIH/NCATS | Institution |
| | | **Time frame: past 36 months** | |
| 2 | Grants or contracts from any entity (if not indicated in item #1 above). | ____NIH | Institution |
| 3 | Royalties or licenses | ____None | |
| 4 | Consulting fees | ____None | |

| | | |
|---|---|---|
| 5 | Payment or honoraria for lectures, presentations, speakers bureaus, manuscript writing or educational events | ____None |
| 6 | Payment for expert testimony | ____None |
| 7 | Support for attending meetings and/or travel | ____None |
| 8 | Patents planned, issued or pending | ____None |
| 9 | Participation on a Data Safety Monitoring Board or Advisory Board | ____None |
| 10 | Leadership or fiduciary role in other board, society, committee or advocacy group, paid or unpaid | ____None |
| 11 | Stock or stock options | ____None |
| 12 | Receipt of equipment, materials, drugs, medical writing, gifts or other services | ____None |
| 13 | Other financial or non-financial interests | ____None |

**Please place an "X" next to the following statement to indicate your agreement:**

**__X_ I certify that I have answered every question and have not altered the wording of any of the questions on this form.**

#### ICMJE DISCLOSURE FORM

**Date:** April 26, 2021
**Your Name:** Hugh Musick
**Manuscript Title:** Rapid implementation of cross-sectional study: Post-acute sequelae of SARS-CoV-2 (PASC) in an ethnically diverse sample in Illinois
**Manuscript number (if known):** Not known

**In the interest of transparency, we ask you to disclose all relationships/activities/interests listed below that are related to the content of your manuscript. "Related" means any relation with for-profit or not-for-profit third parties whose interests may be affected by the content of the manuscript. Disclosure represents a commitment to transparency and does not necessarily indicate a bias. If you are in doubt about whether to list a relationship/activity/interest, it is preferable that you do so.**

**The following questions apply to the author's relationships/activities/interests as they relate to the current manuscript only.**

**The author's relationships/activities/interests should be defined broadly. For example, if your manuscript pertains to the epidemiology of hypertension, you should declare all relationships with manufacturers of antihypertensive medication, even if that medication is not mentioned in the manuscript.**

**In item #1 below, report all support for the work reported in this manuscript without time limit. For all other items, the time frame for disclosure is the past 36 months.**

| | | **Name all entities with whom you have this relationship or indicate none (add rows as needed)** | **Specifications/Comments (e.g., if payments were made to you or to your institution)** |
|---|---|---|---|
| | | **Time frame: Since the initial planning of the work** | |
| 1 | All support for the present manuscript (e.g., funding, provision of study materials, medical writing, article processing charges, etc.) **No time limit for this item.** | ____None | |
| | | **Time frame: past 36 months** | |
| 2 | Grants or contracts from any entity (if not indicated in item #1 above). | ____Patient Centered Outcomes Research Institute | Institution |
| 3 | Royalties or licenses | ____None | |

| | | |
|---|---|---|
| 4 | Consulting fees | ____None |
| 5 | Payment or honoraria for lectures, presentations, speakers bureaus, manuscript writing or educational events | ____None |
| 6 | Payment for expert testimony | ____None |
| 7 | Support for attending meetings and/or travel | ____None |
| 8 | Patents planned, issued or pending | ____None |
| 9 | Participation on a Data Safety Monitoring Board or Advisory Board | ____None |
| 10 | Leadership or fiduciary role in other board, society, committee or advocacy group, paid or unpaid | ____None |
| 11 | Stock or stock options | ____None |
| 12 | Receipt of equipment, materials, drugs, medical writing, gifts or other services | ____None |
| 13 | Other financial or non-financial interests | ____None |

**Please place an "X" next to the following statement to indicate your agreement:**

X **I certify that I have answered every question and have not altered the wording of any of the questions on this form.**

#### ICMJE DISCLOSURE FORM

**Date:** __4/27/21_
**Your Name:** __Richard M. Novak__
**Manuscript Title:** Rapid implementation of cross-sectional study: Post-acute sequelae of SARS-CoV-2 (PASC) in an ethnically diverse sample in Illinois
**Manuscript number (if known):** Not known

**In the interest of transparency, we ask you to disclose all relationships/activities/interests listed below that are related to the content of your manuscript. "Related" means any relation with for-profit or not-for-profit third parties whose interests may be affected by the content of the manuscript. Disclosure represents a commitment to transparency and does not necessarily indicate a bias. If you are in doubt about whether to list a relationship/activity/interest, it is preferable that you do so.**

**The following questions apply to the author's relationships/activities/interests as they relate to the current manuscript only.**

**The author's relationships/activities/interests should be defined broadly. For example, if your manuscript pertains to the epidemiology of hypertension, you should declare all relationships with manufacturers of antihypertensive medication, even if that medication is not mentioned in the manuscript.**

**In item #1 below, report all support for the work reported in this manuscript without time limit. For all other items, the time frame for disclosure is the past 36 months.**

| | | **Name all entities with whom you have this relationship or indicate none (add rows as needed)** | **Specifications/Comments (e.g., if payments were made to you or to your institution)** |
|---|---|---|---|
| | | **Time frame: Since the initial planning of the work** | |
| 1 | All support for the present manuscript (e.g., funding, provision of study materials, medical writing, article processing charges, etc.) **No time limit for this item.** | __x__None | |
| | | **Time frame: past 36 months** | |
| 2 | Grants or contracts from any entity (if not indicated in item #1 above). | __x__None | |
| 3 | Royalties or licenses | _x___None | |
| 4 | Consulting fees | __x__None | |

| | | |
|---|---|---|
| 5 | Payment or honoraria for lectures, presentations, speakers bureaus, manuscript writing or educational events | __x__None |
| 6 | Payment for expert testimony | __x__None |
| 7 | Support for attending meetings and/or travel | _x___None |
| 8 | Patents planned, issued or pending | __x__None |
| 9 | Participation on a Data Safety Monitoring Board or Advisory Board | __x__None |
| 10 | Leadership or fiduciary role in other board, society, committee or advocacy group, paid or unpaid | _x___None |
| 11 | Stock or stock options | __x__None |
| 12 | Receipt of equipment, materials, drugs, medical writing, gifts or other services | __x__None |
| 13 | Other financial or non-financial interests | __x__None |

**Please place an "X" next to the following statement to indicate your agreement:**

**__x_ I certify that I have answered every question and have not altered the wording of any of the questions on this form.**

### ICMJE DISCLOSURE FORM

**Date:** April 25, 2021
**Your Name:** Heather M. Prendergast MD, MS, MPH
**Manuscript Title:** Rapid implementation of cross-sectional study: Post-acute sequelae of SARS-CoV-2 (PASC) in an ethnically diverse sample in Illinois
**Manuscript number (if known):** Not known

**In the interest of transparency, we ask you to disclose all relationships/activities/interests listed below that are related to the content of your manuscript. "Related" means any relation with for-profit or not-for-profit third parties whose interests may be affected by the content of the manuscript. Disclosure represents a commitment to transparency and does not necessarily indicate a bias. If you are in doubt about whether to list a relationship/activity/interest, it is preferable that you do so.**

**The following questions apply to the author's relationships/activities/interests as they relate to the current manuscript only.**

**The author's relationships/activities/interests should be defined broadly. For example, if your manuscript pertains to the epidemiology of hypertension, you should declare all relationships with manufacturers of antihypertensive medication, even if that medication is not mentioned in the manuscript.**

**In item #1 below, report all support for the work reported in this manuscript without time limit. For all other items, the time frame for disclosure is the past 36 months.**

| | | Name all entities with whom you have this relationship or indicate none (add rows as needed) | Specifications/Comments (e.g., if payments were made to you or to your institution) |
|---|---|---|---|
| | | **Time frame: Since the initial planning of the work** | |
| 1 | All support for the present manuscript (e.g., funding, provision of study materials, medical writing, article processing charges, etc.) **No time limit for this item.** | ____None | |
| | | **Time frame: past 36 months** | |
| 2 | Grants or contracts from any entity (if not indicated in item #1 above). | NIH | Institution: NHLBI: Targeting of Uncontrolled Hypertension (TOUCHED) |
| | | Sergey Brin Family Foundation | Institution |
| | | CDC | Institution: National Center for Chronic Disease Prevention and Health Promotion Special Emphasis Panel: Improving Cancer Outcomes through Appropriate ED evaluation (ICARE) |

| | | |
|---|---|---|
| 3 | Royalties or licenses | ____None |
| 4 | Consulting fees | ____None |
| 5 | Payment or honoraria for lectures, presentations, speakers bureaus, manuscript writing or educational events | ____None |
| 6 | Payment for expert testimony | ____None |
| 7 | Support for attending meetings and/or travel | ____None |
| 8 | Patents planned, issued or pending | ____None |
| 9 | Participation on a Data Safety Monitoring Board or Advisory Board | ____None |
| 10 | Leadership or fiduciary role in other board, society, committee or advocacy group, paid or unpaid | Cook County Health & Hospital Board |
| 11 | Stock or stock options | ____None |
| 12 | Receipt of equipment, materials, drugs, medical writing, gifts or other services | ____None |
| 13 | Other financial or non-financial interests | ____None |

**Please place an "X" next to the following statement to indicate your agreement:**

__X_ **I certify that I have answered every question and have not altered the wording of any of the questions on this form.**
